# Comparing palliative care quality between designated and non-designated cancer hospitals: A secondary analysis of bereaved family surveys

**DOI:** 10.64898/2026.05.14.26353232

**Authors:** Satomi Ito, Mitsunori Miyashita, Richi Takahashi, Yoko Nakazawa, Asao Ogawa, Nobuyuki Yotani, Jun Hamano

**Affiliations:** Department of Palliative Nursing, Health Sciences, Tohoku University Graduate School of Medicine, 2-1 Seiryo-machi, Aoba-ku, Sendai, Miyagi 980-8575, Japan; Division of Policy Evaluation, Institute for Cancer Control, National Cancer Center, 5-1-1 Tsukiji, Chuo-ku, Tokyo 104-0045, Japan; Division of Quality Assurance Programs, Institute for Cancer Control, National Cancer Center, 5-1-1 Tsukiji, Chuo-ku, Tokyo 104-0045, Japan; Psycho Oncology Division, Research Center for Innovative Oncology, National Cancer Center Hospital East, 6-5-1 Kashiwanoha, Kashiwa-shi, Chiba 277-8577, Japan; Division of Palliative Medicine, National Center for Child Health and Development, 2-10-1 Okura, Setagaya-ku, Tokyo 157-8535, Japan; Department of Palliative and Supportive Care, Institute of Medicine, University of Tsukuba, 1-1-1 Tennoudai, Tsukuba, Ibaraki 305-8575, Japan

**Keywords:** palliative care, end-of-life, evaluation, nationwide survey, cancer

## Abstract

**Background:** The quality of palliative care in non-designated cancer hospitals, where approximately 70% of deaths of patients with cancer occur, remains unevaluated. This study aimed to clarify the quality of palliative care in these hospitals by comparing patient characteristics and evaluating the quality of palliative care provided by bereaved families.

**Methods:** A questionnaire survey was conducted among bereaved family members of patients with cancer who died in 2018 at designated and non-designated cancer hospitals (excluding palliative care units). We compared the two groups regarding patient and bereaved family characteristics, quality assessment of palliative care (including Memorial Symptom Assessment Scale [MSAS]), care satisfaction, and the presence of end-of-life discussions.

**Results:** In total, 27,944 bereaved family members agreed to participate. The mean age at death was 73.2 (±11.9) and 79.7 (±10.9) years for designated and non-designated cancer hospitals, respectively (p < 0.001, Effect Size [ES] = 0.55). The mean MSAS total score (symptom intensity) was significantly higher for designated cancer hospitals than for non-designated cancer hospitals, even after adjusting for patient characteristics (p < 0.001, ES = 0.39). Conversely, the mean adjusted overall satisfaction was significantly higher in non-designated cancer hospitals (p < 0.001, ES = 0.21) than in designated cancer hospitals.

**Conclusions:** Non-designated cancer hospitals had older and less symptomatic patients than designated cancer hospitals. However, there was no significant clinical difference in the quality of palliative care, as assessed by the bereaved families.

## Introduction

Evaluating palliative care is important for maintaining and improving palliative care quality [1]. However, patients receiving palliative care are physically and mentally vulnerable, making it difficult for them to participate in surveys [2]. Therefore, as an alternative, evaluation of the quality of palliative care from the perspective of bereaved families has been implemented worldwide [3–7].

The quality and delivery of palliative care at designated cancer hospitals are well-documented [8,9]. Approximately 400 designated cancer hospitals in Japan, operating under the Cancer Control Act, are central to developing standardized, high-quality comprehensive cancer care [10]. These hospitals must have Palliative Care Consultation Teams comprising full-time physicians, nurses, psychiatrists, and pharmacists to provide high-quality palliative care. Palliative cancer-care services in designated cancer hospitals significantly developed between 2008 and 2018 [9]. However, palliative care services in community hospitals remain insufficient. Furthermore, the quality of palliative care in non-designated cancer hospitals, where approximately 70% of hospital cancer deaths occur, remains unevaluated [11].

Therefore, this study aimed to clarify the quality of palliative care in non-designated cancer hospitals by comparing patient characteristics and care quality assessments by bereaved families with those of designated cancer hospitals.

### Patients and methods

This study was a secondary analysis of data from a previous survey of bereaved families whose relatives died in hospitals, excluding palliative care units. The Institutional Review Board of the National Cancer Center in Japan approved this study’s scientific and ethical validity (Research project no.: 2023–127). The names of designated cancer hospitals and hospitals with palliative care physicians were obtained from open data [12].

The study population comprised patients with cancer who died in 2018. Bereaved family members served as proxies for assessment. Participants were identified through stratified random sampling based on the 2018 Vital Statistics Survey of Death Lists from the Ministry of Health, Labor, and Welfare, stratified by place of death.

A mailed questionnaire survey was conducted. The research team mailed survey request forms, letters of intent, survey instruments, return envelopes, and pens to family members 13–25 months post-bereavement. Respondents were asked to return their completed surveys in unmarked, self-addressed envelopes within 2 weeks of receiving the survey form. Approximately 1 month after the initial mailing, a reminder was sent to non-respondents. The survey was conducted between March and May 2020.

This study was conducted and reported in accordance with the STROBE (Strengthening the Reporting of Observational Studies in Epidemiology) guidelines.

#### Measurement

The primary endpoint was the difference in palliative care quality. Secondary endpoints included differences in end-of-life discussion rates and patient/bereaved family characteristics.

#### Care Evaluation Scale (CES)

The CES, a 28-item measure of medical care structure and process at the time of death [13], was used with 13 items in this study. Bereaved families rated palliative care quality on a 7-point Likert scale (1 = improvement is highly necessary, 6 = improvement is not necessary, 0 = unknown). Total scores ranged from 0 to 78, with higher scores indicating greater satisfaction.

#### Good Death Inventory (GDI)

The GDI, an 18-item measure, quantifies the achievement of the desired death in patients with cancer from the perspective of the bereaved family [13]. Families rated the quality of medical care on a 7-point Likert scale ranging from 1 (absolutely disagree) to 7 (absolutely agree). Total scores ranged from 18 to 126, with higher scores indicating greater satisfaction with the quality of palliative care.

#### Memorial Symptom Assessment Scale (MSAS)

MSAS is a 32-item measure to quantify physical symptoms common to patients with cancer [14]. Eleven of these were used in this study. Bereaved families were asked to rate the intensity of symptoms on a 6-point Likert scale ranging from 1 (none) to 5 (very severe), with 0 = unknown. Total scores ranged from 0 to 55, with higher scores indicating higher levels of distress.

#### Satisfaction

Satisfaction, a single-item measure, assessed overall satisfaction with medical care received at the place of death. Bereaved families rated the medical care their patient received on a 6-point Likert scale ranging from 1 (very unsatisfactory) to 6 (very satisfied).

#### End-of-life discussion

End-of-life discussions were assessed using a 5-item measure, quantifying preferences and discussion status regarding the end-of-life care location and medical care. Bereaved families were asked to rate discussion status on a 4-point Likert scale ranging from 1 (not implemented) to 4 (implemented).

#### Participants’ characteristics

Additional survey items included patient background (such as age at death, length of time until death, pre-death activities of daily living [ADL] status, and presence of dementia) and bereaved family background (such as age, sex, relationship with the patient, employment status, and pre-death mental and physical health).

#### Statistical analysis

Responses with missing Care Evaluation Scale (CES), Good Death Inventory (GDI), or MSAS items, and those from participants whose relatives died in palliative care units were excluded. A descriptive analysis of demographic characteristics was performed by dividing the patients into two groups: those who died at designated versus non-designated cancer hospitals. We compared demographic characteristics between the patients’ death locations using the Student’s t test and the chi-square test. Effect Size (ES) was calculated using Cohen’s d and Cramer’s V to assess the magnitude of the clinical differences. We compared the total CES, GDI, and MSAS scores and satisfaction scores between the patients’ death locations using the Student’s t test and Cohen’s d. We then conducted a multiple regression analysis with CES, GDI, and MSAS total scores and satisfaction as objective variables, and patient characteristics as explanatory variables to control for confounding factors. From the results, we compared the differences in the respective adjusted means using the Student’s t test and Cohen’s d. End-of-life discussion implementation was converted to binary variables: ‘not implemented’ (including ‘probably not implemented’) and ‘implemented’ (including ‘probably implemented’). We compared the status of end-of-life discussions between patients’ death locations using the Student’s t test and Cramer’s V. Statistical significance was set at p < 0.05. All analyses were performed using SAS® software (version 9.4; SAS Institute, Cary, NC, USA).

## Results

Of 310,543 eligible patients, questionnaires were sent to 61,100 families, yielding 27,944 valid responses, with 13,995 included in the analysis (Fig 1).

**Fig 1.**
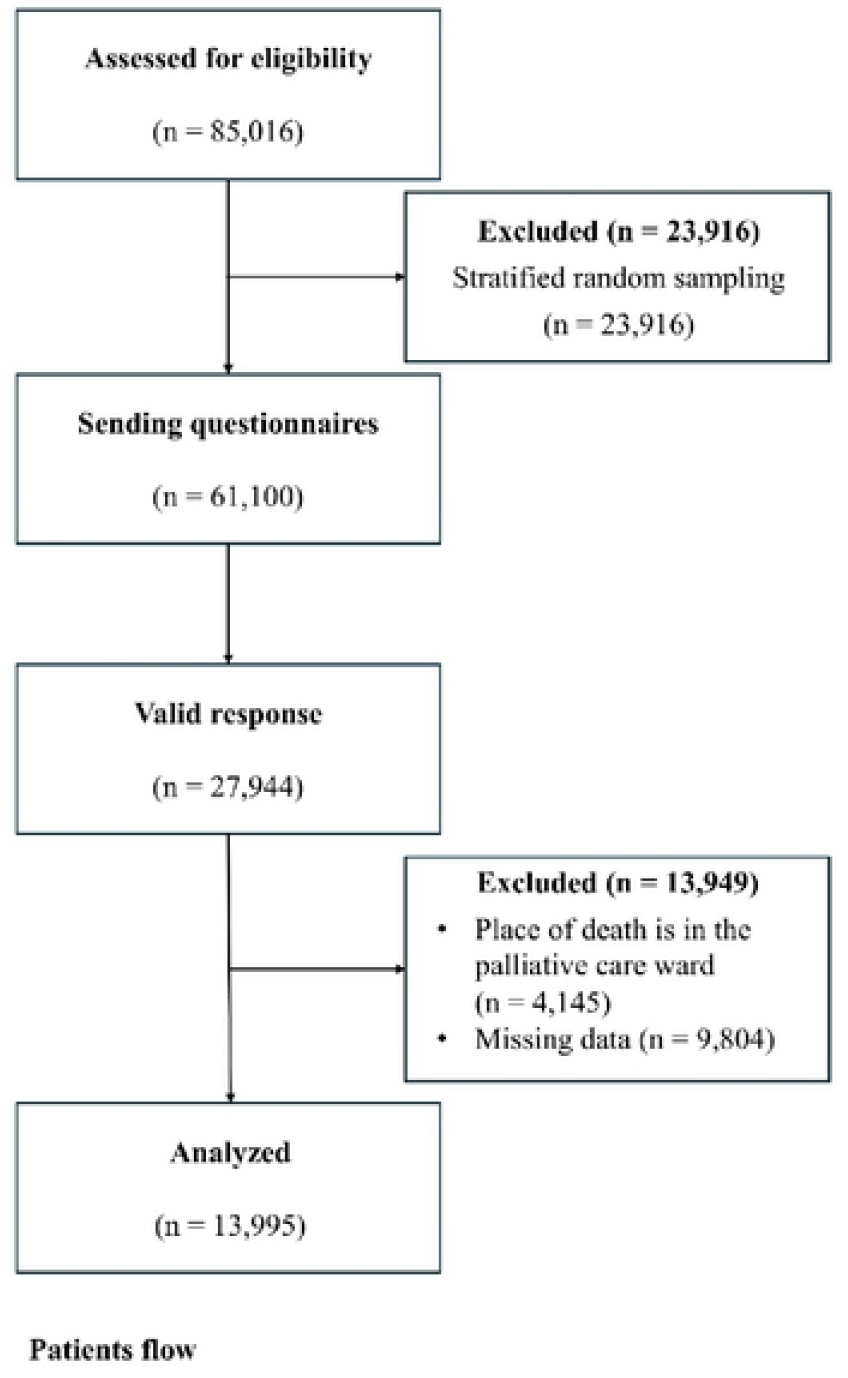
Patient flow. Abbreviations: PCU, palliative care unit.

### Patient characteristics

A total of 4,821 patients died in designated cancer hospitals and 9,174 in non-designated hospitals. The patient characteristics revealed notable differences between hospital types (Table 1). Patients in non-designated cancer hospitals were significantly older than those in designated hospitals, with a mean age of 79.7 vs 73.2 years. This age difference had a moderate effect size (p < 0.001, ES = 0.55), suggesting a meaningful distinction. Moreover, patients in non-designated hospitals showed greater impairment in Activities of Daily Living (ADL), communication ability, and a higher prevalence of dementia. These findings highlight that non-designated hospitals tend to care for older, frailer patients.

**Table 1.**
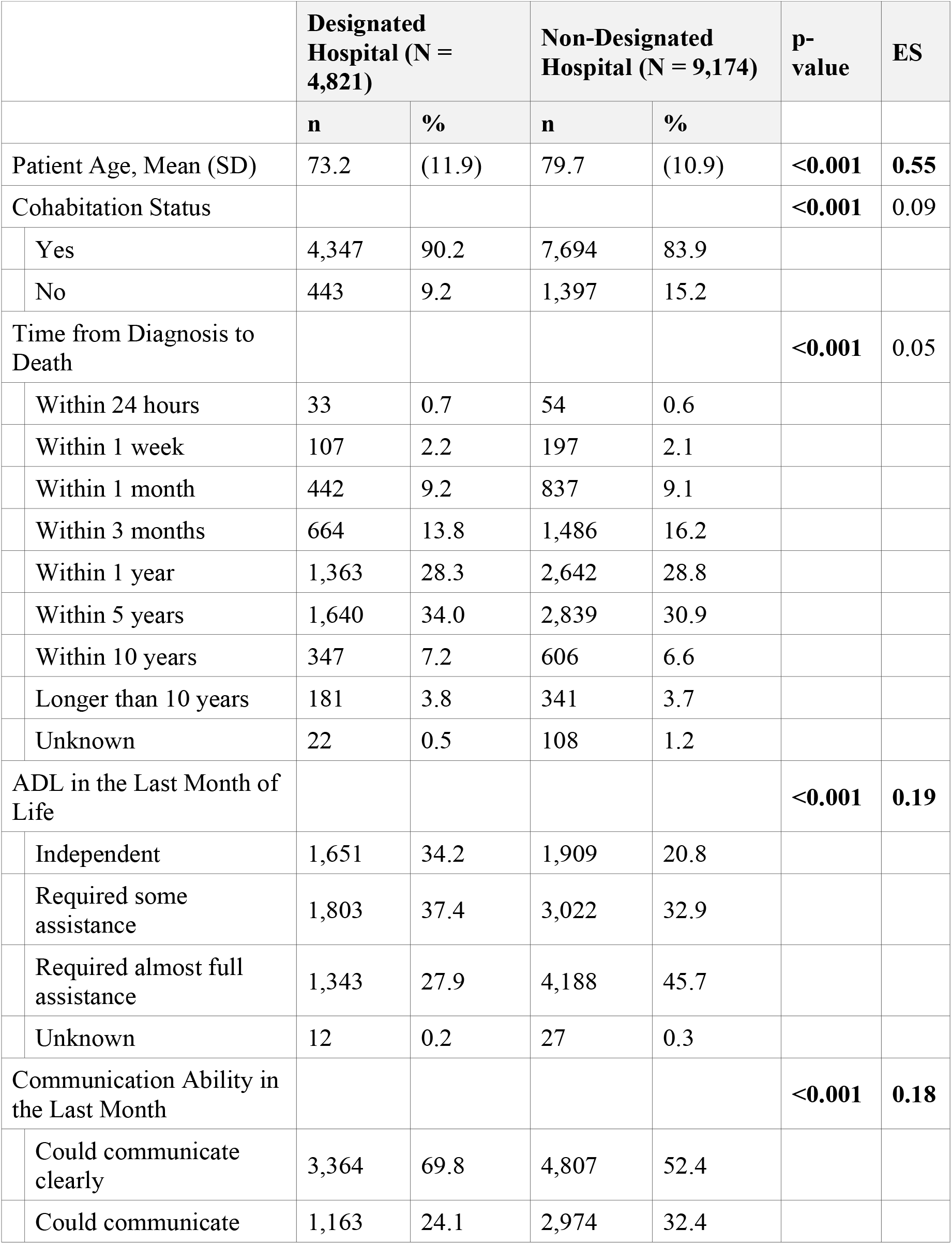
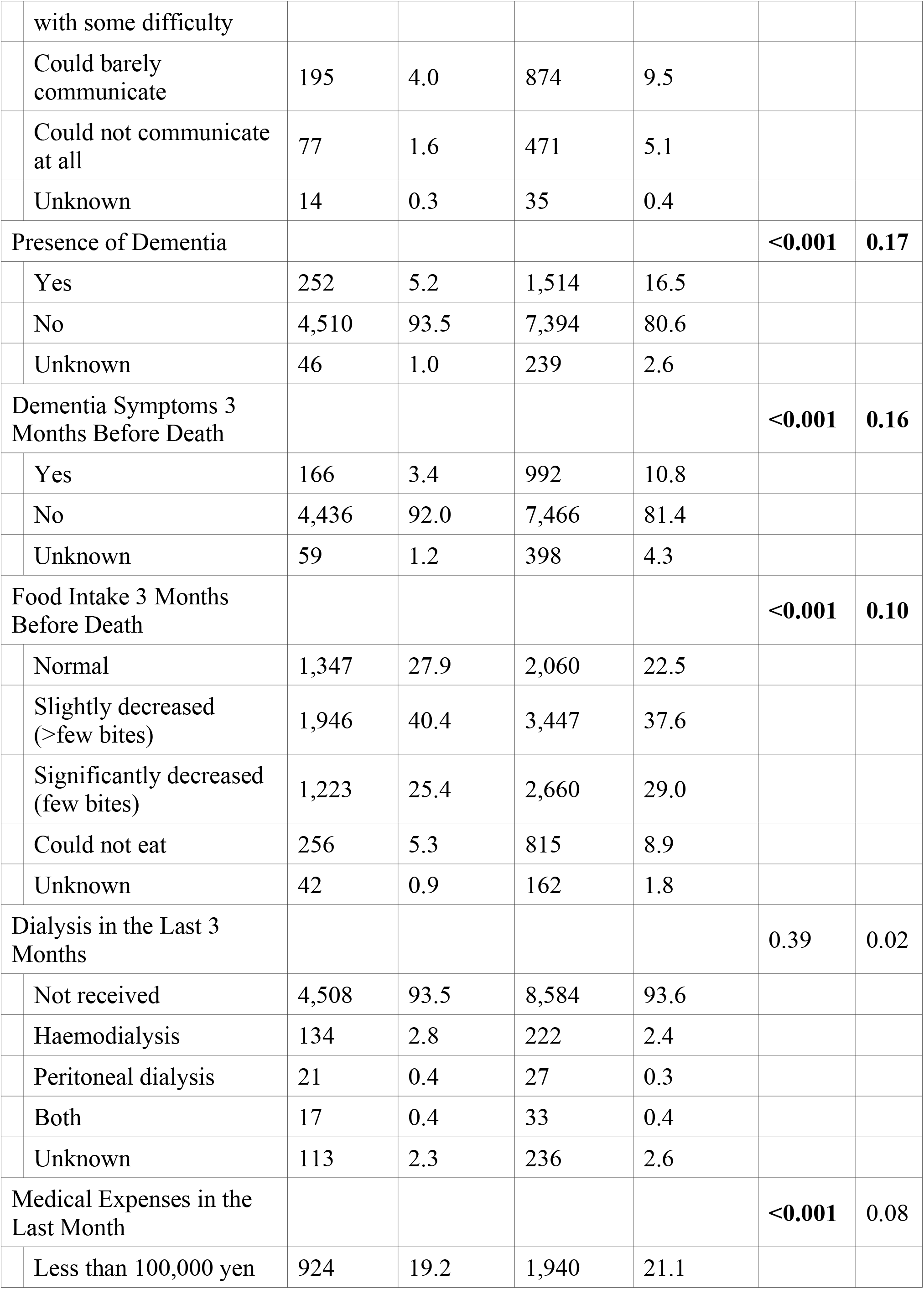
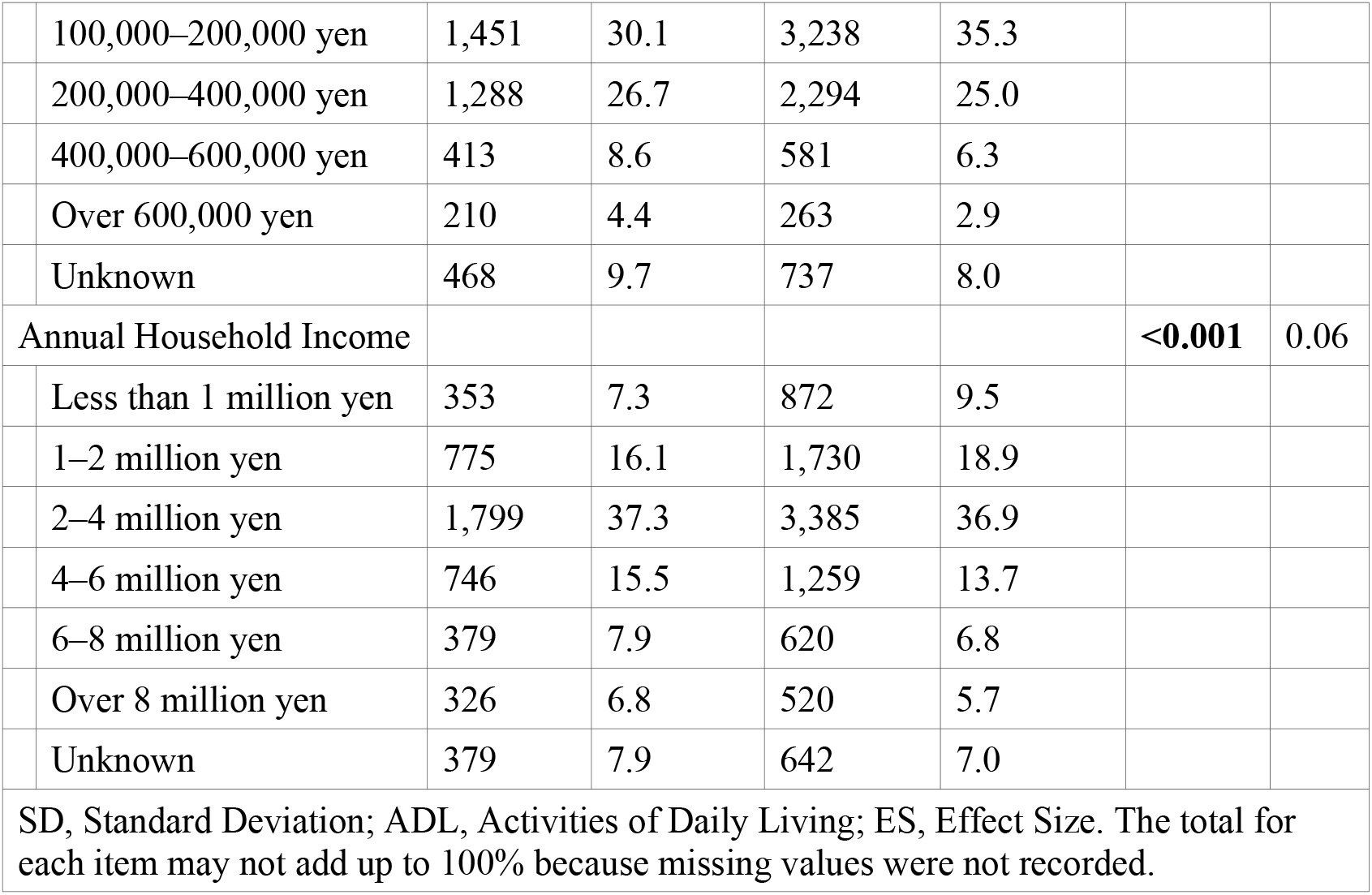
Patient characteristics.

| Table 1. Patient characteristics |  |  |  |  |  |  |
| --- | --- | --- | --- | --- | --- | --- |
|  | Designated Hospital (N = 4,821) |  | Non-Designated Hospital (N = 9,174) |  | p-value | ES |
|  | n | % | n | % |  |  |
| Patient Age, Mean (SD) | 73.2 | (11.9) | 79.7 | (10.9) | **<0.001** | **0.55** |
| Cohabitation Status |  |  |  |  | **<0.001** | 0.09 |
| Yes | 4,347 | 90.2 | 7,694 | 83.9 |  |  |
| No | 443 | 9.2 | 1,397 | 15.2 |  |  |
| Time from Diagnosis to Death |  |  |  |  | **<0.001** | 0.05 |
| Within 24 hours | 33 | 0.7 | 54 | 0.6 |  |  |
| Within 1 week | 107 | 2.2 | 197 | 2.1 |  |  |
| Within 1 month | 442 | 9.2 | 837 | 9.1 |  |  |
| Within 3 months | 664 | 13.8 | 1,486 | 16.2 |  |  |
| Within 1 year | 1,363 | 28.3 | 2,642 | 28.8 |  |  |
| Within 5 years | 1,640 | 34.0 | 2,839 | 30.9 |  |  |
| Within 10 years | 347 | 7.2 | 606 | 6.6 |  |  |
| Longer than 10 years | 181 | 3.8 | 341 | 3.7 |  |  |
| Unknown | 22 | 0.5 | 108 | 1.2 |  |  |
| ADL in the Last Month of Life |  |  |  |  | **<0.001** | **0.19** |
| Independent | 1,651 | 34.2 | 1,909 | 20.8 |  |  |
| Required some assistance | 1,803 | 37.4 | 3,022 | 32.9 |  |  |
| Required almost full assistance | 1,343 | 27.9 | 4,188 | 45.7 |  |  |
| Unknown | 12 | 0.2 | 27 | 0.3 |  |  |
| Communication Ability in the Last Month |  |  |  |  | **<0.001** | **0.18** |
| Could communicate clearly | 3,364 | 69.8 | 4,807 | 52.4 |  |  |
| Could communicate | 1,163 | 24.1 | 2,974 | 32.4 |  |  |

|  |  |  |  |  |  |  |  |
| --- | --- | --- | --- | --- | --- | --- | --- |
|  | with some difficulty |  |  |  |  |  |  |
|  | Could barely communicate | 195 | 4.0 | 874 | 9.5 |  |  |
|  | Could not communicate at all | 77 | 1.6 | 471 | 5.1 |  |  |
|  | Unknown | 14 | 0.3 | 35 | 0.4 |  |  |
| Presence of Dementia |  |  |  |  |  | **<0.001** | **0.17** |
|  | Yes | 252 | 5.2 | 1,514 | 16.5 |  |  |
|  | No | 4,510 | 93.5 | 7,394 | 80.6 |  |  |
|  | Unknown | 46 | 1.0 | 239 | 2.6 |  |  |
| Dementia Symptoms 3 Months Before Death |  |  |  |  |  | **<0.001** | **0.16** |
|  | Yes | 166 | 3.4 | 992 | 10.8 |  |  |
|  | No | 4,436 | 92.0 | 7,466 | 81.4 |  |  |
|  | Unknown | 59 | 1.2 | 398 | 4.3 |  |  |
| Food Intake 3 Months Before Death |  |  |  |  |  | **<0.001** | **0.10** |
|  | Normal | 1,347 | 27.9 | 2,060 | 22.5 |  |  |
|  | Slightly decreased (>few bites) | 1,946 | 40.4 | 3,447 | 37.6 |  |  |
|  | Significantly decreased (few bites) | 1,223 | 25.4 | 2,660 | 29.0 |  |  |
|  | Could not eat | 256 | 5.3 | 815 | 8.9 |  |  |
|  | Unknown | 42 | 0.9 | 162 | 1.8 |  |  |
| Dialysis in the Last 3 Months |  |  |  |  |  | 0.39 | 0.02 |
|  | Not received | 4,508 | 93.5 | 8,584 | 93.6 |  |  |
|  | Haemodialysis | 134 | 2.8 | 222 | 2.4 |  |  |
|  | Peritoneal dialysis | 21 | 0.4 | 27 | 0.3 |  |  |
|  | Both | 17 | 0.4 | 33 | 0.4 |  |  |
|  | Unknown | 113 | 2.3 | 236 | 2.6 |  |  |
| Medical Expenses in the Last Month |  |  |  |  |  | **<0.001** | 0.08 |
|  | Less than 100,000 yen | 924 | 19.2 | 1,940 | 21.1 |  |  |
|  | 100,000–200,000 yen | 1,451 | 30.1 | 3,238 | 35.3 |  |  |
|  | 200,000–400,000 yen | 1,288 | 26.7 | 2,294 | 25.0 |  |  |
|  | 400,000–600,000 yen | 413 | 8.6 | 581 | 6.3 |  |  |
|  | Over 600,000 yen | 210 | 4.4 | 263 | 2.9 |  |  |
|  | Unknown | 468 | 9.7 | 737 | 8.0 |  |  |
| Annual Household Income |  |  |  |  |  | **<0.001** | 0.06 |
|  | Less than 1 million yen | 353 | 7.3 | 872 | 9.5 |  |  |
|  | 1–2 million yen | 775 | 16.1 | 1,730 | 18.9 |  |  |
|  | 2–4 million yen | 1,799 | 37.3 | 3,385 | 36.9 |  |  |
|  | 4–6 million yen | 746 | 15.5 | 1,259 | 13.7 |  |  |
|  | 6–8 million yen | 379 | 7.9 | 620 | 6.8 |  |  |
|  | Over 8 million yen | 326 | 6.8 | 520 | 5.7 |  |  |
|  | Unknown | 379 | 7.9 | 642 | 7.0 |  |  |
| SD, Standard Deviation; ADL, Activities of Daily Living; ES, Effect Size. The total for each item may not add up to 100% because missing values were not recorded. |  |  |  |  |  |  |  |

### Bereaved family characteristics

The characteristics of bereaved family members showed no clinically significant differences were observed across any items (Table 2). For example, the mean age of bereaved families was 61.7 (SD = 12.3) and 62.9 years (SD = 11.6) in designated and non-designated hospitals, respectively (p < 0.001, ES = 0.10). Although there were statistically significant differences in sex, and relationship to the patient, all effect sizes were below 0.2, indicating negligible clinical relevance. This suggests a broadly similar demographic profile of family respondents across both hospital types.

**Table 2.** Characteristics of the bereaved families of deceased patients.

|  |  |  |  |
| --- | --- | --- | --- |
|  | Designated | Non-Designated | p-value |

|  |  | Hospital (N = 4,821) |  | Hospital (N = 9,174) |  |  |
| --- | --- | --- | --- | --- | --- | --- |
|  |  | n | % | n | % | ES |
| Bereaved family age, Mean (SD) |  | 61.7 | (12.3) | 62.9 | (11.6) | <0.001; 0.10 |
| Sex of bereaved family |  |  |  |  |  | <0.001; 0.04 |
|  | Male | 1,803 | 37.4 | 3,805 | 41.5 |  |
|  | Female | 2,993 | 62.1 | 5,315 | 57.9 |  |
| Relationship |  |  |  |  |  | <0.001; 0.16 |
|  | Spouse | 2,310 | 47.9 | 2,932 | 32.0 |  |
|  | Child | 1,835 | 38.1 | 4,628 | 50.4 |  |
|  | Son-in-law / Daughter-in-law | 322 | 6.7 | 817 | 8.9 |  |
|  | Parent | 95 | 2.0 | 194 | 2.1 |  |
|  | Other relatives | 209 | 4.3 | 481 | 5.2 |  |
|  | Other | 22 | 0.5 | 64 | 0.7 |  |
| Mutual understanding (could understand each other's feelings) |  |  |  |  |  | <0.001; 0.04 |
|  | Strongly disagree | 133 | 2.8 | 270 | 2.9 |  |
|  | Disagree | 299 | 6.2 | 709 | 7.7 |  |
|  | Slightly agree | 1,477 | 30.6 | 2,920 | 31.8 |  |
|  | Agree | 2,881 | 59.8 | 5,206 | 56.7 |  |
| Employment during last week before patient's death |  |  |  |  |  | <0.001; 0.08 |
|  | Not working | 1,961 | 40.7 | 3,637 | 39.6 |  |
|  | Was working but quit | 230 | 4.8 | 442 | 4.8 |  |
|  | Took a leave | 504 | 10.5 | 590 | 6.4 |  |
|  | Working part-time | 579 | 12.0 | 1,119 | 12.2 |  |
|  | Working full-time | 1,487 | 30.8 | 3,256 | 35.5 |  |
| Extent of care provided in last week before |  |  |  |  |  | <0.001; 0.09 |
| death |  |  |  |  |  |  |
|  | No care provided | 421 | 8.7 | 1,025 | 11.2 |  |
|  | 1–3 days/week | 690 | 14.3 | 1,833 | 20.0 |  |
|  | 4–6 days/week | 623 | 12.9 | 1,207 | 13.2 |  |
|  | Every day | 3,037 | 63.0 | 5,009 | 54.6 |  |
| Physical health condition during last week before death |  |  |  |  |  | 0.32;<br>0.02 |
|  | Very poor | 130 | 2.7 | 293 | 3.2 |  |
|  | Poor | 676 | 14.0 | 1,308 | 14.3 |  |
|  | Fair | 2,527 | 52.4 | 4,816 | 52.5 |  |
|  | Good | 1,461 | 30.3 | 2,700 | 29.4 |  |
| Mental health condition during last week before death |  |  |  |  |  | <0.001;<br>0.07 |
|  | Very poor | 546 | 11.3 | 832 | 9.1 |  |
|  | Poor | 1,607 | 33.3 | 2,642 | 28.8 |  |
|  | Fair | 2,051 | 42.5 | 4,337 | 47.3 |  |
|  | Good | 582 | 12.1 | 1,289 | 14.1 |  |
| SD, Standard Deviation; ES, Effect Size. The total for each item may not add up to 100% because missing values were not recorded. p-value and ES are shown as "p-value; ES" in combined cells. |  |  |  |  |  |  |

### Palliative care quality evaluation

After adjusting for patient characteristics, MSAS total scores were significantly higher in designated hospitals (ES = 0.39) than in non-designated hospitals (Table 3). This suggests that patients in these hospitals experienced more intense symptom burdens. Conversely, overall satisfaction with medical care, as rated by bereaved family members, was slightly higher in non-designated cancer hospitals. Although this difference was statistically significant (p < 0.001), the effect size (ES = 0.21) was small, indicating a limited clinical impact. CES and GDI scores did not show meaningful clinical differences after adjustment. These results suggest that while designated hospitals manage more symptomatic patients, the perceived quality of palliative care by families does not differ substantially between hospital types.

**Table 3.** Evaluation of the quality of palliative care.

| **Table 3. Evaluation of the quality of palliative care** |  |  |  |  |  |  |  |
| --- | --- | --- | --- | --- | --- | --- | --- |
|  |  | **Designated Hospital (N = 4,821)** |  | **Non-Designated Hospital (N = 9,174)** |  | **p-value** | **ES** |
|  |  | **Mean** | **SD** | **Mean** | **SD** |  |  |
| CES Total | Before adjustment | 44.3 | 9.6 | 43.7 | 9.8 | <0.001 | 0.06 |
|  | After adjustment | 44.0 | 2.2 | 43.9 | 2.2 | 0.56 | 0.01 |
| GDI Total | Before adjustment | 65.6 | 21.5 | 63.3 | 22.9 | <0.001 | 0.10 |
|  | After adjustment | 65.3 | 9.3 | 63.7 | 10.0 | <0.001 | 0.16 |
| MSAS Total | Before adjustment | 26.1 | 11.0 | 24.2 | 11.2 | <0.001 | 0.17 |
|  | After adjustment | 26.0 | 3.9 | 24.4 | 4.2 | <0.001 | 0.39 |
| Satisfaction with Medical Care | Before adjustment | 4.4 | 1.4 | 4.4 | 1.4 | 0.60 | 0.01 |
|  | After adjustment | 4.3 | 0.2 | 4.4 | 0.2 | <0.001 | 0.21 |
| SD, Standard Deviation; CES, Care Evaluation Scale; GDI, Good Death Inventory; MSAS, Memorial Symptom Assessment Scale; ES, Effect Size. |  |  |  |  |  |  |  |

### End-of-life discussions

Table 4 shows the status of end-of-life discussions regarding preferences regarding care location and medical treatment preferences. End-of-life discussions between patients, family members, and healthcare providers were similar across hospital types. While statistical differences were observed in some items, such as discussions about place of care and resuscitation, none reached the threshold for clinical relevance (ES < 0.2). These findings imply that the opportunity and extent of such discussions were comparably implemented regardless of hospital designation.

**Table 4.** Discussions regarding end-of-life care and desired place of care.

|  |  |  |  |  |  |  |
| --- | --- | --- | --- | --- | --- | --- |
|  | **Designated**<br>**Hospital (N =**<br>**4,821)** |  | **Non-Designated**<br>**Hospital (N =**<br>**9,174)** |  | **p-**<br>**value** | **ES** |
|  | **n** | **%** | **n** | **%** |  |  |
| Discussion about desired place of care (Patient–Primary doctor) |  |  |  |  | <0.001 | 0.03 |
| Was able to have the discussion | 1,393 | 28.9 | 2,723 | 29.7 |  |  |
| Was not able to have the discussion | 1,155 | 24.0 | 2,575 | 28.1 |  |  |
| Discussion about resuscitation procedures (Patient–Primary doctor) |  |  |  |  | 0.11 | 0.02 |
| Was able to have the discussion | 1,653 | 34.3 | 3,162 | 34.5 |  |  |
| Was not able to have the discussion | 1,127 | 23.4 | 1,995 | 21.7 |  |  |
| Creation of documents regarding resuscitation procedures |  |  |  |  | 0.46 | 0.01 |
| Was able to create the documents | 603 | 12.5 | 1,199 | 13.1 |  |  |
| Was not able to create the documents | 3,393 | 70.4 | 6,479 | 70.6 |  |  |
| Discussion about place of care and resuscitation procedures (Patient–Bereaved family) |  |  |  |  | <0.001 | 0.03 |
| Was able to have the discussion | 1,852 | 38.4 | 3,713 | 40.5 |  |  |
| Was not able to have the discussion | 2,252 | 46.7 | 4,063 | 44.3 |  |  |
| Discussion about resuscitation procedures (Bereaved family–Primary doctor) |  |  |  |  | <0.001 | 0.04 |
| Was able to have the discussion | 2,685 | 55.7 | 5,466 | 59.6 |  |  |
| Was not able to have the discussion | 1,289 | 26.7 | 2,164 | 23.6 |  |  |
| ES, Effect Size. The total for each item may not add up to 100% because missing values were not recorded. |  |  |  |  |  |  |

## Discussion

To the best of our knowledge, this study is the first to clarify differences in palliative care quality using data from bereaved family members of patients with cancer who died at designated and non-designated cancer hospitals. The main finding of this study is that there were no clinically significant differences in the quality of palliative care between designated and non-designated cancer hospitals, as assessed using CES, GDI, MSAS, and satisfaction scores. This indicates that, from the perspective of bereaved family members, palliative care quality is relatively comparable regardless of hospital designation. This suggests that standardization efforts, such as generalist palliative care training programs, may have contributed to levelling the quality of care across institutions. As a secondary finding, we observed demographic differences in the patient population: those in non-designated hospitals were older and exhibited greater functional decline, including lower ADL and higher prevalence of dementia. These demographic factors may reflect differences in patient referral patterns or hospital roles within local care systems.

This study aimed to clarify the quality of palliative care between designated and non-designated cancer hospitals. Considering this objective, our discussion emphasizes that while non-designated hospitals cared for older and less symptomatic patients, the perceived quality of palliative care as evaluated by bereaved families was not significantly different between hospital types. This finding contradicts our assumption, based on previous findings, that non-designated cancer hospitals provide inadequate palliative care services, including palliative care team interventions, compared with those provided in designated cancer hospitals [9]. This highlights that hospital designation status alone may not be a sufficient indicator of palliative care quality. Thus, patient characteristics and care structures must be jointly considered in evaluating quality of care.

Furthermore, although designated hospitals are generally better equipped for palliative care, the absence of perceived quality differences may reflect evolving capabilities in non-designated hospitals. One factor contributing to this may be physician training programs such as the Palliative care Emphasis program on symptom management and Assessment for Continuous medical Education (PEACE) program [15]. This national initiative, aimed at all physicians, provides foundational education in palliative care across hospital types. As of 2018, 113,907 physicians had completed the program [16]. However, we cannot confirm the extent of PEACE participation specifically within non-designated hospitals. Nevertheless, its broad implementation may have helped standardize basic palliative care delivery.

This study has some limitations. First, the results of this study, obtained from secondary analysis of proxy evaluations of bereaved families, may not always align fully with patient evaluations. However, obtaining direct evaluations from patients in the terminal phase is often difficult, and the validity of our method is supported by previous studies which also used proxy evaluations by bereaved families [17,18]. Second, the transfer status of patients in their last month of life was unknown. Although the study investigated the quality of palliative care and patient characteristics from 1 month to 1 week before death, there may have been discrepancies between the actual place of care and the place of death. Third, with a 54% response rate, non-respondents may have provided lower ratings for palliative care quality and end-of-life discussions. When interpreting the results, the possibility of overestimation owing to non-response bias must be considered. Future studies should leverage data such as medical claims to facilitate comprehensive comparisons of palliative care services, improving the evaluation of hospital and patient characteristics. Furthermore, our classification of hospitals as designated or non-designated cancer hospitals was based solely on the place of death recorded in official statistics. This method does not capture whether patients received most of their treatment at the facility where they ultimately died. In addition, we could not assess the involvement of specialist palliative care teams in the end-of-life phase. These uncertainties may have influenced the bereaved families’ perception of care quality and should be acknowledged as key limitations of this study.

In conclusion, this study found that non-designated cancer hospitals had older and less symptomatic patients than designated cancer hospitals; however, bereaved families reported no significant clinical difference in palliative care quality.

## Supporting information

**S1 Checklist**. STROBE checklist for observational studies.

## Funding

This work was funded by MHLW Research for Promotion of Cancer Control Program Grant Number 23EA1021. The funders had no role in study design, data collection and analysis, decision to publish, or preparation of the manuscript.

## Acknowledgments

This work was supported by JST SPRING, Grant Number JPMJSP2114 and Advanced Graduate Program for Future Medicine and Health Care, Tohoku University.

## Author contributions

Conceptualization: Satomi Ito, Mitsunori Miyashita, Richi Takahashi, Yoko Nakazawa, Asao

Ogawa, Nobuyuki Yotani, Jun Hamano.

Data curation: Richi Takahashi, Yoko Nakazawa.

Formal analysis: Satomi Ito.

Writing – original draft: Satomi Ito.

Writing – review & editing: Mitsunori Miyashita, Richi Takahashi, Yoko Nakazawa, Asao

Ogawa, Nobuyuki Yotani, Jun Hamano.

## Competing interests

The authors declare no competing interests.

## Data availability

Data cannot be shared publicly because they contain personal information that could compromise research participant privacy. Data are available from the Institutional Review Board of the National Cancer Center in Japan (contact via) for researchers who meet the criteria for access to confidential data.

## Ethics statement

The Institutional Review Board of the National Cancer Center in Japan approved this study’s scientific and ethical validity (Research project no.: 2023–127). Informed consent was obtained from all participants.

## Notes

### Competing Interest Statement

The authors have declared no competing interest.

### Funding Statement

Yes

### Author Declarations

This study was approved by the Institutional Review Board of the National Cancer Center Japan (Research project no.: 2023–127). Written informed consent was obtained from all participants by return of the completed mailed questionnaire.

## References

1. Teno JM. Measuring end-of-life care outcomes retrospectively. J Palliat Med. 2005;8 Suppl 1:S42–9.

2. Addington-Hall JM, MacDonald LD, Anderson HR, Chamberlain J, Freeling P, Bland JM, et al. Randomised controlled trial of effects of coordinating care for terminally ill cancer patients. BMJ. 1992;305(6865):1317–22.

3. Addington-Hall J, McCarthy M. Regional Study of Care for the Dying: methods and sample characteristics. Palliat Med. 1995;9(1):27–35.

4. Lynn J, Teno JM, Phillips RS, Wu AW, Desbiens N, Harrold J, et al. Perceptions by family members of the dying experience of older and seriously ill patients. Ann Intern Med. 1997;126(2):97–106.

5. Costantini M, Beccaro M, Merlo F, ISDOC Study Group. The last three months of life of Italian cancer patients. Palliat Med. 2005;19(8):628–38.

6. Seamark DA, Williams S, Hall M, Lawrence CJ, Gilbert J. Dying from cancer in community hospitals or a hospice: closest lay carers’ perceptions. Br J Gen Pract. 1998;48(431):1317–21.

7. Addington-Hall J, Altmann D, McCarthy M. Which terminally ill cancer patients receive hospice in-patient care? Soc Sci Med. 1998;46(8):1011–6.

8. Maeda I, Tsuneto S, Miyashita M, Morita T, Umeda M, Motoyama M, et al. Progressive development and enhancement of palliative care services in Japan: nationwide surveys of designated cancer care hospitals for three consecutive years. J Pain Symptom Manage. 2014;48(3):364–73.

9. Nakazawa Y, Kato M, Miyashita M, Morita T, Ogawa A, Kizawa Y. Growth and challenges in hospital palliative cancer care services: an analysis of nationwide surveys over a decade in Japan. J Pain Symptom Manage. 2021;61(6):1155–64.

10. Ministry of Health Labour and Welfare. Basic plan to promote cancer control programs [Internet]. 2007 [cited 2025 Jan 9]. Available from: http://www.mhlw.go.jp/bunya/kenkou/gan_keikaku.html

11. National Cancer Center. Current status report data for all designated cancer hospitals, etc [Internet]. Cancer Information Service. [cited 2025 Mar 11]. Available from: https://hospdb.ganjoho.jp/download_file/4128/0

12. Ministry of Health, Labour and Welfare, Japan. Hospitals Designated as Cancer Treatment Centers [Internet]. [cited 2025 Jul 20]. Available from: https://www.mhlw.go.jp/stf/seisakunitsuite/bunya/kenkou_iryou/kenkou/gan/gan_byoin.html

13. Miyashita M, Aoyama M, Nakahata M, Yamada Y, Abe M, Yanagihara K, et al. Development of the Care Evaluation Scale version 2.0: a modified version of a measure for bereaved family members to evaluate the structure and process of palliative care for cancer patients. BMC Palliat Care. 2017;16(1):8.

14. Portenoy RK, Thaler HT, Kornblith AB, McCarthy Lepore J, Friedlander-Klar H, Kiyasu E, et al. The Memorial Symptom Assessment Scale: an instrument for the evaluation of symptom prevalence, characteristics and distress. Eur J Cancer. 1994;30(9):1326–36.

15. Yamamoto R, Kizawa Y, Nakazawa Y, Ohde S, Tetsumi S, Miyashita M. Outcome evaluation of the palliative care emphasis program on symptom management and assessment for continuous medical education: nationwide physician education project for primary palliative care in Japan. J Palliat Med. 2015;18(1):45–9.

16. Takamiya Y, Ozawa M, Shimizu T, editors. Hospice and Palliative Care White Paper 2025. Tokyo: Seikaiysa; 2025. p. 58–67.

17. Masukawa K, Aoyama M, Morita T, Kizawa Y, Tsuneto S, Shima Y, et al. The Japan hospice and palliative evaluation study 4: a cross-sectional questionnaire survey. BMC Palliat Care. 2018;17(1):115.

18. Office for National Statistics, England NHS. National Survey of Bereaved People, 2015 [Internet]. UK Data Service; 2016. Available from: http://dx.doi.org/10.5255/UKDA-SN-7979-1

